# Mental and social health of children and adolescents with pre-existing mental or somatic problems during the COVID-19 pandemic lockdown

**DOI:** 10.1101/2020.12.15.20248237

**Authors:** Josjan Zijlmans, Lorynn Teela, Hanneke van Ewijk, Helen Klip, Malindi van der Mheen, Hyun Ruisch, Michiel A. J. Luijten, Maud M. van Muilekom, Kim J. Oostrom, Jan Buitelaar, Pieter J. Hoekstra, Ramón Lindauer, Arne Popma, Wouter Staal, Robert Vermeiren, Hedy A. van Oers, Lotte Haverman, Tinca J. C. Polderman

**Affiliations:** Amsterdam University Medical Center, Vrije Universiteit Amsterdam, Department of Child and Adolescent Psychiatry & Psychosocial Care, Amsterdam Public Health, Amsterdam, The Netherlands; Emma Children’s Hospital, Amsterdam UMC, University of Amsterdam, Child and Adolescent Psychiatry & Psychosocial Care, Amsterdam Reproduction and Development, Amsterdam Public Health, Amsterdam, The Netherlands; Curium-LUMC, Leiden University Medical Center, Leiden, The Netherlands; Karakter Child and Adolescent Psychiatry University Centre, Nijmegen, The Netherlands; Amsterdam University Medical Center, University of Amsterdam, Department of Child and Adolescent Psychiatry, Amsterdam, The Netherlands; Levvel, Academic Center for Child and Adolescent Psychiatry, Amsterdam, The Netherlands; University of Groningen, University Medical Center Groningen, Department of Child and Adolescent Psychiatry, Groningen, The Netherlands; Amsterdam University Medical Center, Vrije Universiteit Amsterdam, Epidemiology and Data Science, Amsterdam Public Health, Amsterdam, The Netherlands; Department of Cognitive Neuroscience, Donders Institute for Brain, Cognition and Behaviour, Radboudumc, Nijmegen, The Netherlands

## Abstract

**Background:** The COVID-19 lockdown increases psychological problems in children and adolescents from the general population. Here we investigate the mental and social health during the COVID-19 lockdown in children and adolescents with pre-existing mental or somatic problems.

**Method:** We included participants (8-18 years) from a psychiatric (N = 249) and pediatric (N = 90) sample, and compared them to a general population sample (N = 844). Measures were assessed during the first lockdown (April-May 2020) in the Netherlands. Main outcome measures were Patient-Reported Outcomes Measurement Information System (PROMIS®) domains: Global Health, Peer Relationships, Anxiety, Depressive Symptoms, Anger, and Sleep-Related Impairment. Additionally, socio-demographic variables, COVID-19-related questions, changes in atmosphere at home from a parent and child perspective, and children’s experiences of lockdown regulations were assessed.

**Results:** On all measures except Global Health, the pediatric sample reported least problems. The psychiatric sample reported significantly more problems than the general population sample on all measures except for Anxiety and Peer Relationships. Having a COVID-19 affected friend/relative and a COVID-19 related change in work situation negatively moderated outcome, but not in the samples with pre-existing problems. All parents reported significant decreases in atmosphere at home, as did children from the general population.

**Conclusion:** We observed significant differences in mental and social health between three child and adolescent samples during the COVID-19 pandemic lockdown and identified COVID-19-related factors influencing mental and social health. Our findings contribute to current and future policies during pandemic related lockdowns regarding the mental and social health of children and adolescents.

## Introduction

It is well known that large scale disasters have great impact on the well-being of the general population. Whether human-induced (e.g., the World Trade Center attacks), natural (e.g., hurricanes), or technological (e.g., Chernobyl nuclear reactor accident), disasters are accompanied by an increase in a wide scale of mental and behavioral disorders ^1^. Pandemics specifically can lead to increased levels of anxiety, depression, and post-traumatic stress ^2,3^. The COVID-19 pandemic is a major game changer in the life of many individuals across the world. Important direct consequences that highly affect social life are regulations imposed by governments, such as closing schools, and leisure and sports facilities, and obligating social distancing in the public space. During COVID-19, increased rates of anxiety and depression, and decreased psychological well-being have been reported in the adult general population ^4^, and adult psychiatric patients show worsening of their symptoms ^5^.

Empirical studies on mental and social health during the COVID-19 pandemic in children and adolescents are limited ^6^, though several cross-sectional Chinese studies have been published. These studies have shown that 20-40% of Chinese children and adolescents reported psychological problems during the COVID-19 pandemic, especially symptoms of anxiety, depression, and post-traumatic stress ^7–9^. Very recently, a Dutch study similarly showed that the mental and social health of children and adolescents from the general population had deteriorated since the COVID-19 lockdown ^10^. Growing up in a single-parent family, having more than three children in the family, a negative change in work situation of parents due to COVID-19 regulations, and having a friend or relative infected with COVID-19 were factors associated with more mental and social health problems during the COVID-19 lockdown.

The current study extends these efforts by investigating mental and social health during the Dutch COVID-19 lockdown in vulnerable populations, namely children and adolescents with pre-existing mental problems or with a (chronic) somatic condition. Since children with mental problems and chronic somatic conditions are known to be at risk for psychosocial problems ^11,12^, it is important to examine mental and social health during the COVID-19 lockdown ^13,14^. Psychosocial problems can arise by the fact that activities of (mental) support programs have been disrupted due to the COVID-19 regulations ^15^, and treatments of children and their families suddenly transitioned from in-person therapy to online therapy, which may in some cases affect the quality of their treatment ^16^. To date, empirical studies of the mental and social health of these vulnerable populations during the COVID-19 lockdown are still sparse.

As of yet, few empirical studies have investigated the mental and social effects of COVID-19 related regulations in children with pre-existing mental problems (psychiatric population) and results are mixed. One study in children with autism spectrum disorder (ASD) reported increases in irritability, hyperactivity, inappropriate speech, and speech quality from before to during the pandemic ^17^. Another study in a neuropsychiatric child and adolescent sample reported increases in obsessive-compulsive problems, post-traumatic stress, and thought problems ^18^. On the other hand, a French study investigated the well-being of 533 children and adolescents with self-reported Attention Deficit/Hyperactivity Disorder (ADHD) during the COVID-19 pandemic ^19^. As reported by their parents, 65.3% of these children showed no changes in well-being or were doing better than before the pandemic, suggesting that more than half of the children and adolescents with ADHD experience stability or improvement of their well-being during the COVID-19 lockdown. Still, one third of children reported lower wellbeing, indicating substantial variation in responses. Similarly, anecdotal evidence from child and adolescent clinical practice suggested that for some children and families, COVID-19 lockdown-induced changes reduced sensory exposure and daily stress (e.g., school related stress), and subsequently seemed to reduce mental problems and even improved well-being in some children, while for other children negative outcomes (e.g., increased stress levels, reduced well-being and mental health) are to be expected ^20^.

The literature on the consequences of the COVID-19 lockdown in children with a (chronic) somatic condition (pediatric population) is also limited. A recent study has shown that psychosocial stress in children and adolescents with cancer did not increase during the first months of the COVID-19 pandemic in the Netherlands ^21^. Additionally, adolescents with cystic fibrosis reported lower anxiety levels during the COVID-19 pandemic compared to their healthy peers ^22^. These studies suggest that children with a (chronic) somatic condition, despite their vulnerability, may be less susceptible to negative consequences of the COVID-19 pandemic. However, a recent study in a large Hongkong population (N 29,202 families) showed that children with chronic diseases and children with parents reporting mental problems scored higher on emotional and behavioral problems, and decreased peer relations ^23^. In sum, research in vulnerable groups of children and adolescents during the COVID-19 pandemic is currently scarce, and findings hint at positive as well as negative consequences on mental and social health.

The aim of the current study was to assess the mental and social health of a broad population of children with pre-existing conditions (psychiatric or pediatric) during the first COVID-19 lockdown in the Netherlands and to compare their global health, peer relationships, symptom levels of anxiety, depression and anger, and sleep-related impairment to the general population of children and adolescents. We examined potential associated factors, including COVID-19 specific factors such as having an affected friend or relative. In addition, we described changes in home atmosphere due to the first Dutch COVID-19 lockdown and report qualitative data on children’s experiences of the COVID-19 regulations.

## Methods

### The first COVID-19 lockdown in the Netherlands

On March 15^th^ 2020, the first COVID-19 lockdown came into practice in the Netherlands. From that moment on, all schools and child day-care facilities were closed (unless one or both parents had a profession that was classified as essential), as well as sport and leisure facilities, bars, and restaurants. Adults were advised to work from home as much as possible. People over the age of 12 were advised to keep social distance (1.5 meter) and traveling was discouraged. However, it was still permitted to go outside, to receive three guests at home, and young children (<12 years) could play outside with their friends.

### Procedure

All families were invited to participate during the first COVID-19 lockdown in The Netherlands (between the end of April and early May 2020). During our measurement window, no large changes in Dutch COVID-19 regulations occurred. Parents were approached via email and children were subsequently approached by their parents. If parents and children were willing to participate, the parents completed a sociodemographic questionnaire and COVID-19-related questions online. The children also completed online COVID-19-related questions and several validated Patient-Reported Outcome Measures (PROMs). We collected data through research websites of the KLIK PROM portal developed specifically for each group (www.corona-survey.nl, www.corona.hetklikt.nu, and www.corona-studie.nl). The completion time for children and parents together was approximately 15 minutes. All children and parents provided informed consent and the study was approved by the Medical Ethics Committee of the Amsterdam UMC.

### Participants

We included independent samples of children and adolescents (8-18 years) from child and adolescent psychiatric centers and a children’s hospital and compared these to a sample from the general population.

The psychiatric sample consists of children receiving psychiatric care for varying problems (e.g., autism, depression, ADHD) at one of four tertiary academic child and adolescent psychiatric centers in the Netherlands: Accare, de Bascule/Levvel, Curium, and Karakter. The four centers cover child and adolescent psychiatric care in the northern, western, and eastern part of the Netherlands. We approached parents through email between April 30^th^ and May 12^th^, 2020. In total, we invited 5,615 children and their parents to participate.

The pediatric sample consists of children with a (chronic) somatic condition (e.g., juvenile idiopathic arthritis, endocrinological diseases, cystic fibrosis) who are under treatment in the Emma Children’s Hospital Amsterdam UMC. Children who already complete PROMs as standard part of their care were included. We approached parents via email in the first week of May 2020. In total, we invited 1,240 children and their parents to participate.

For the general population sample, data was collected via an independent online research agency ‘Panel Inzicht’ in April 2020 (see also ^10^). Parents were invited until 1,000 responses were reached.

### Socio-demographic and COVID-related measures

The socio-demographic questionnaire was completed by parents and comprised questions about themselves (region, country of birth, education level, marital status, and number of children) and their child (age, gender). The COVID-19-related questions for parents concerned whether or not the parent or partner reported a negative change in work situation due to COVID-19 (i.e., loss of income, reduced number of working hours, unemployment) and whether or not a friend or relative was infected with COVID-19. Additionally, parents completed two questions regarding the atmosphere at home before and during the first Dutch COVID-19 lockdown on a visual analogue scale (range 0-100): “How did you experience the atmosphere at home before the Corona regulations?” and “How do you experience the atmosphere at home now?”.

The COVID-19-related questions for children also assessed the atmosphere at home: “How did you experience atmosphere at home before the schools were closed?” and “How do you experience the atmosphere at home now?”. Finally, we asked an open-ended question: “How are the Corona-regulations for you?”.

### Mental and social health outcomes

We administered six Dutch Patient-Reported Outcomes Measurement Information System (PROMIS®) pediatric measures: Scale V1.0 - Global health ^24^, CAT V2.0 - Peer Relationships ^25^, CAT V2.0 - Anxiety ^26^, CAT V2.0 - Depressive Symptoms ^26^, Scale V2.0 - Anger ^27^, and CAT V1.0 - Sleep-related Impairment ^28^. These measures have been selected by the American Psychiatric Association (APA) as level-2 assessment measures for monitoring and evaluating psychiatric disorders from the DSM-V. PROMIS measures use a 7-day recall period and items are scored on a five-point Likert scale. All items range from ‘never’ to ‘(almost) always’, except for Global Health, where response categories differ for each item (e.g., ‘excellent’ to ‘poor’). Total scores are calculated by transforming item scores into a T-score ranging from 0 to 100, with a mean of 50 and a standard deviation of 10 in the U.S. general population (for our purposes, we compared to representative Dutch data; see data analysis). For all measures, higher scores represent more of the construct. For Anger (9 items) and Global Health (7+2 items) we used the total scales. For the other item banks, we used Computerized Adaptive Testing (CAT), in which items are presented based on the responses to previously administered items. This results in a reliable total score while administering fewer items than regular questionnaires ^29^.

### Data analysis

We used the Statistical Package for Social Sciences (SPSS) version 26.0 for statistical analyses. First, we characterized participants of each sample using descriptive analysis (means and percentages).

Second, we performed analyses of covariance (ANCOVA) to assess differences in PROMIS outcome measures between samples. We corrected for age, sex, and parental country of birth as these are characteristics known to influence mental and social outcomes ^11,30,31^. We performed Bonferroni-corrected post-hoc t-tests to determine which samples differed significantly from each other. Next, we tested whether there were differences in proportions of children reporting severe symptoms or poor functioning between samples. ‘Severe’ symptoms and ‘poor’ functioning were defined as 1.5 SD above or below the mean T-score of an independent and representative pre-COVID-19 Dutch sample ^32,33^, except for Peer Relations, where 2 SD was used as the cut-off value (see www.HealthMeasures.net).

Third, to determine which variables were associated with the PROMIS measures during COVID-19, we repeated the ANCOVA over all groups and included pertinent independent variables: marital status, region, number of children in the family, parental educational level, change in work situation due to COVID-19, and infected relative/friend with COVID-19. As the proportion of the participants from the general population is high, we repeated the ANCOVA without the general population sample to explore whether effects are mainly driven by this group.

Fourth, we performed paired samples T-tests in each group to investigate changes in atmosphere at home from before to during COVID-19 as reported by children and parents. In addition, we performed an ANCOVA to assess whether the changes in atmosphere differed between samples, correcting for age and sex.

Finally, LT and HAvO qualitatively analyzed the children’s answers to the open-ended question “How are the corona-regulations for you?” using the method for thematic analysis in psychology ^34^. Answers were categorized into positive, neutral, or negative experiences and clustered into main themes. Themes were ranked based on the number of times mentioned by children (most to least mentioned). We analyzed answers from all groups together but explored differences between groups.

## Results

### Sociodemographic characteristics

Sociodemographic characteristics of all samples are presented in Table 1. In the psychiatric sample 249 children participated (response rate: 4.4%). The mean age was 12.8 years, 56.2% was male, 17.7% had an infected relative/friend, and 18.9% of their parents experienced a negative change in work situation due to COVID-19.

**Table 1.** Socio-demographic characteristics of participants per group.

|  | Child/adolescent<br>psychiatric<br>sample | Pediatric<br>sample | General<br>population<br>sample |
| --- | --- | --- | --- |
| *N* | 249 | 90 | 844 |
| Mean age in years (SD) | 12.8 (2.9) | 12.9 (3.3) | 13.4 (2.8) |
|  | % | % | % |
| Sex (male) | 56.2 | 55.6 | 47.4 |
| Region<sup>^</sup> |  |  |  |
| North | 30.1 | 3.3 | 12.6 |
| East | 26.1 | 13.3 | 22.2 |
| South | 1.2 | 1.1 | 25.0 |
| West | 42.6 | 82.2 | 40.3 |
| Number of children in<br>family |  |  |  |
| 1 child | 16.9 | 18.2 | 25.5 |
| 2 children | 53.4 | 51.1 | 46.6 |
| 3 children or more | 29.7 | 30.7 | 27.9 |
| Country of birth parents |  |  |  |
| Netherlands (both<br>parents) | 83.9 | 83.1 | 88.2 |
| Foreign country<br>(at least one parent) | 16.1 | 16.9 | 11.8 |
| Marital status parents |  |  |  |
| Two-parent family | 76.8 | 81.1 | 82.0 |
| Single-parent family | 23.2 | 18.9 | 18.0 |
| Educational level<br>parents° |  |  |  |
| Low | 9.2 | 6.7 | 9.0 |
| Intermediate | 43.8 | 45.6 | 51.8 |
| High | 47.0 | 47.8 | 39.2 |
| COVID-19-specific<br>variables |  |  |  |
| Infected relative/friend<br>(yes) | 17.7 | 33.3 | 23.7 |
| Negative change in<br>work<br>situation (yes) | 18.9 | 21.1 | 26.2 |

**Note.** Due to missing values, number of respondents differs slightly across socio-demographic variables, minimal Ns are 815, 243, and 88. ^Region: North = Groningen, Friesland, Drenthe, East = Overijssel, Gelderland, Flevoland, South = Zeeland, Noord-Brabant, Limburg, West = Utrecht, Noord-Holland, Zuid-Holland. °Educational level parents, Low; primary, lower vocational education, lower and middle general secondary education; Intermediate: middle vocational education, higher secondary education, pre-university education; High: higher vocational education, university.

In the pediatric sample 90 children participated (response rate: 7.1%). The mean age was 12.9 years, 55.6% was male, 33.3% had an infected relative/friend, and 21.1% of their parents had a negative change in work situation due to COVID-19.

In the general population sample 844 children participated (response rate: 8.4%). The mean age was 13.4 years, 47.4% was male, 23.7% had an infected relative/friend, and 26.2% of their parents experienced a negative change in work situation due to COVID-19.

### Between group differences in mental and social health

We found significant differences on all PROMIS measures between the three groups (p < 0.01, η^2^ = 0.03 – 0.04) after controlling for the covariates age, sex, and parental country of birth (see Table 2). The psychiatric sample reported worst mental and social health and the pediatric sample reported best mental and social health, whereas the general population had scores in between. Post-hoc t-tests revealed that the psychiatric sample reported significantly worse scores than the general population (p < 0.05) on the Global Health (ΔM = -3.31, CI = -4.58; -2.05), Depressive Symptoms (ΔM = 2.03, CI = 0.51; 3.56), Anger (ΔM = 3.24, CI = 1.75; 4.74), and Sleep-Related Impairment (ΔM = 2.97, CI = 1.41; 4.53) measures.

**Table 2.**
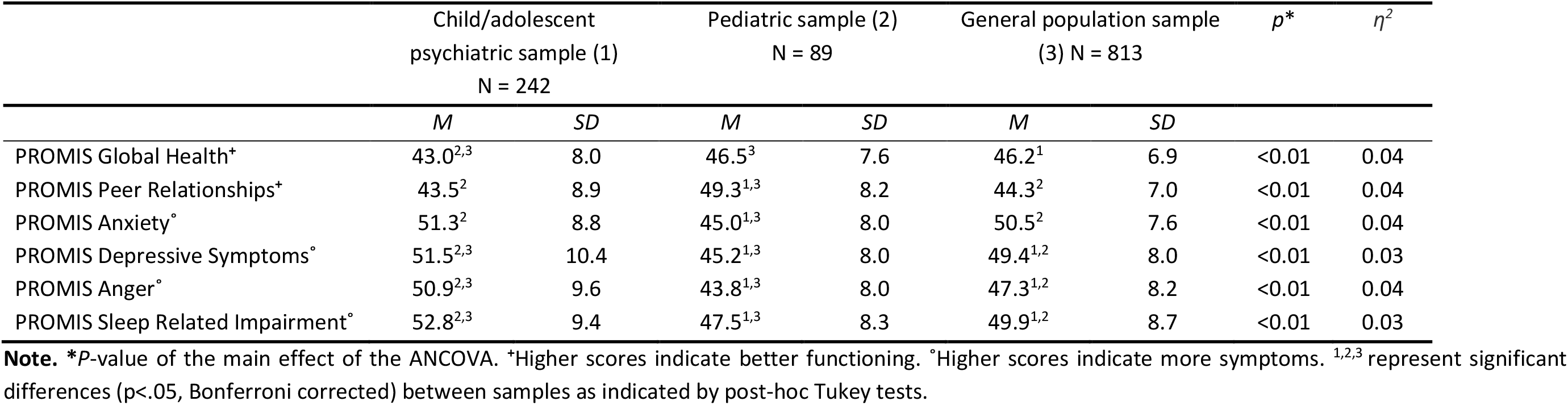
Mean PROMIS T-scores in the different samples during COVID-19, corrected by age, sex, and parental country of birth.

| | Child/adolescent<br>psychiatric sample (1)<br>N = 242 | | Pediatric sample (2)<br>N = 89 | | General population sample<br>(3) N = 813 | | *p* * | $\eta^2$ |
| --- | --- | --- | --- | --- | --- | --- | --- | --- |
|  | *M* | *SD* | *M* | *SD* | *M* | *SD* |  |  |
| PROMIS Global Health<sup>+</sup> | 43.0<sup>2,3</sup> | 8.0 | 46.5<sup>3</sup> | 7.6 | 46.2<sup>1</sup> | 6.9 | <0.01 | 0.04 |
| PROMIS Peer Relationships<sup>+</sup> | 43.5<sup>2</sup> | 8.9 | 49.3<sup>1,3</sup> | 8.2 | 44.3<sup>2</sup> | 7.0 | <0.01 | 0.04 |
| PROMIS Anxiety<sup>°</sup> | 51.3<sup>2</sup> | 8.8 | 45.0<sup>1,3</sup> | 8.0 | 50.5<sup>2</sup> | 7.6 | <0.01 | 0.04 |
| PROMIS Depressive Symptoms<sup>°</sup> | 51.5<sup>2,3</sup> | 10.4 | 45.2<sup>1,3</sup> | 8.0 | 49.4<sup>1,2</sup> | 8.0 | <0.01 | 0.03 |
| PROMIS Anger<sup>°</sup> | 50.9<sup>2,3</sup> | 9.6 | 43.8<sup>1,3</sup> | 8.0 | 47.3<sup>1,2</sup> | 8.2 | <0.01 | 0.04 |
| PROMIS Sleep Related Impairment<sup>°</sup> | 52.8<sup>2,3</sup> | 9.4 | 47.5<sup>1,3</sup> | 8.3 | 49.9<sup>1,2</sup> | 8.7 | <0.01 | 0.03 |

**Note.** \**P*-value of the main effect of the ANCOVA. <sup>+</sup>Higher scores indicate better functioning. <sup>°</sup>Higher scores indicate more symptoms. <sup>1,2,3</sup> represent significant differences (*p*<.05, Bonferroni corrected) between samples as indicated by post-hoc Tukey tests.

The pediatric sample reported significantly better scores than the general population sample (p < 0.05) on Peer Relationships (ΔM = 5.09, CI = 3.06; 7.12), Anxiety (ΔM = -5.67, CI = -7.79; -3.55), Depressive Symptoms (ΔM = -4.23, CI = -6.52; -1.94), Anger (ΔM = -3.72, CI = -5.97; -1.46), and Sleep-Related Impairment (ΔM = -2.44; CI = -4.79; -0.08).

The psychiatric sample reported significantly worse scores than the pediatric sample (p < 0.05) on all six PROMIS domains: Global Health (ΔM = -3.53, CI = -5.62; -1.39), Peer Relationships (ΔM = - 5.87, CI = -8.12; -3.62), Anxiety (ΔM = 6.20, CI = 3.86; 8.55), Depressive Symptoms (ΔM = 6.26; CI = 3.72; 8.80), Anger (ΔM = 6.96, CI = 4.46; 9.45), and Sleep-Related Impairment (ΔM = 5.40, CI = 2.80; 8.01).

Likewise, the percentages of severe/poor scores were highest in the psychiatric sample (range = 5.2 – 22.1%) and lowest in the pediatric sample (range = 0 – 7.8%; see Figure 1).

**Figure 1.**
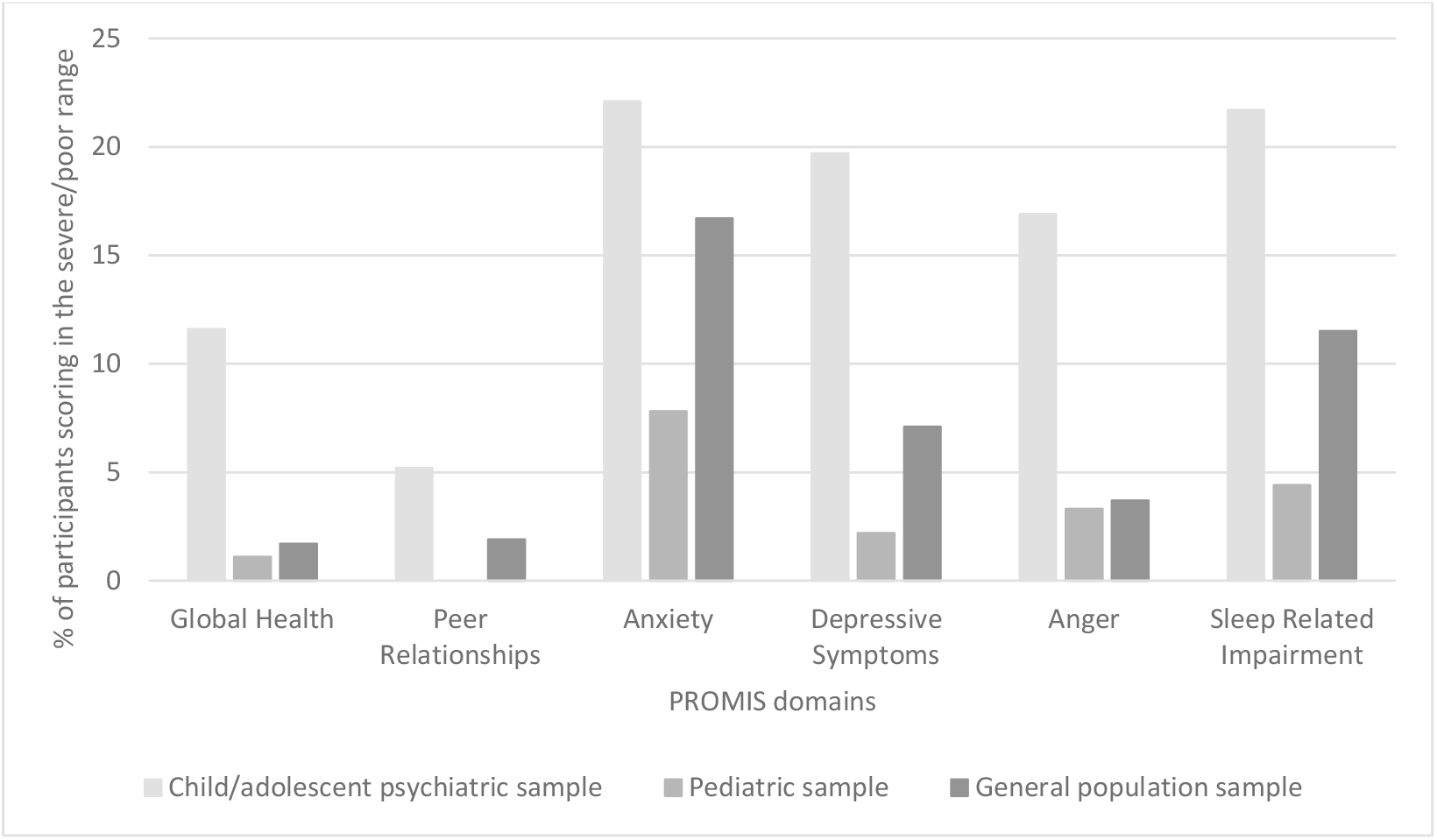
Percentage of participants scoring in the severe/poor range (>1.5 SD)* on PROMIS domains for all samples. **Note**. *For Peer Relationships a cut-off of 2 SD was used, following the HealthMeasures guidelines.

### Variables associated with mental and social health during COVID-19

In Table 3, we report the ANCOVA regression coefficients of the independent variables including all three groups. Higher age was significantly associated (*p* < .01) with lower Global Health (b = -0.21, CI = -0.35; -0.06), higher Anxiety (b = -0.28, CI = -0.44; -0.11), and lower Anger (b = -0.49, CI = -0.66; - 0.32). Male gender was significantly associated with higher Global Health (b = 1.19, CI = 0.36; 2.02), lower Peer Relationships (b = -1.11, CI = -2.00; -0.22), lower Anxiety (b = -1.04, CI = -1.95; -0.12), lower Depressive Symptoms (b = -1.55, CI = -2.55; -0.55), and lower Sleep-related Impairment (b = -1.37; CI = -2.39; -0.35). Living in the northern region of the Netherlands was significantly (*p* < .05) associated with lower scores on Depressive Symptoms (b = -2.00; CI = -3.51; 0.48). Being from a single-parent family was significantly associated (*p* < .05) with lower Global health (b = -2.14, CI = -3.21; -1.08) and higher Anxiety (b = 1.30, CI = 0.13; 2.48). Having three children or more within the household was significantly associated (*p* < .05) with higher Anger (b = 1.77, CI = 0.38; 3.15). For each outcome measure, the group effect remained.

**Table 3.** Unstandardized regression coefficients of independent variables of the ANCOVA for mental and social health outcomes.

|  | Global health | Peer relationships | Anxiety | Depressive symptoms | Anger | Sleep-Related Impairment |
| --- | --- | --- | --- | --- | --- | --- |
| Predictors | B | B | B | B | B | B |
| Age | -0.21** | 0.02 | -0.28* | -0.04 | -0.49** | 0.00 |
| Male sex | 1.20* | -1.11* | -1.04* | -1.55* | -0.12 | -1.37** |
| Region |  |  |  |  |  |  |
| West | Ref | Ref | Ref | Ref | Ref | Ref |
| North | 0.26 | 0.65 | -1.28 | -2.00* | -1.36 | -0.84 |
| East | -0.21 | 0.06 | 0.67 | -0.08 | 0.08 | 0.84 |
| South | -0.61 | -0.07 | -0.00 | -0.68 | -0.61 | -0.68 |
| Foreign country of birth parents | -0.97 | 0.07 | 0.52 | 0.40 | 0.29 | 1.35 |
| Educational level parents° |  |  |  |  |  |  |
| Low | Ref | Ref | Ref | Ref | Ref | Ref |
| Intermediate | -0.50 | 0.62 | 1.17 | 2.02 | 1.88 | 1.64 |
| High | 0.73 | 1.00 | 1.02 | 1.84 | 1.71 | 0.63 |
| Single-parent family | -2.14** | -1.09 | 1.30* | 0.74 | 0.33 | 1.31 |
| Number of children in family |  |  |  |  |  |  |
| 1 child | Ref | Ref | Ref | Ref | Ref | Ref |
| 2 children | 0.08 | -0.69 | -0.17 | -0.03 | 0.93 | 0.31 |
| 3 or more children | 0.81 | 0.15 | -0.34 | -0.02 | 1.77* | 0.48 |
| Negative change in<br>work situation | -0.47 | -0.22 | 2.11** | 1.72** | 1.17 | 2.01** |
| Infected<br>relative/friend with<br>COVID-19 | 0.74 | 0.02 | 1.44** | 0.97 | -0.12 | 0.74 |

**Note.** *B* = Unstandardized regression coefficient of multivariable linear regression model. \*\*p-value <.01, \*p-value <.05. ^Region: North = Groningen, Friesland, Drenthe, East = Overijssel, Gelderland, Flevoland, South = Zeeland, Noord-Brabant, Limburg, West = Utrecht, Noord-Holland, Zuid-Holland. °Educational level parents divided into three categories: Low = primary, lower vocational education, lower and middle general secondary education; Intermediate = middle vocational education, higher secondary education, pre-university education; High = higher vocational education, university

Regarding COVID-19-specific variables, a negative change in parental work situation was significantly associated (p < 0.01) with more Anxiety (b = 2.11; CI = 1.05; 3.18), more Depressive Symptoms (b = 1.72, CI = 0.56; 2.88), more Anger (b = 1.17, CI = 0.03; 2.31), and more Sleep-Related Impairment (b = 2.01, CI = 0.82; 3.20). A parent having a friend or relative infected with COVID-19 was significantly associated (p < 0.01) with a higher Anxiety score (b = 1.44, CI = 0.35; 2.53).

Finally, we performed the ANCOVA analyses again in the psychiatric and pediatric groups only to explore whether effects are driven mainly by the general population sample. We found higher age to be significantly associated (*p* < .01) with lower Global Health (b = -0.51; CI = -0.81; -0.22) and lower Anger (b = -0.48, CI = -0.83; -0.12). Male sex was significantly associated (*p* < .05) with higher Global Health (b = 2.38, CI = 0.65; 4.11), lower Anxiety (b = -2.27, CI = -4.20; -0.33), and lower Depressive Symptoms (b = -3.24, CI = -5.45; -1.03). Living in the northern region of the Netherlands was significantly (*p* < .05) associated with lower Anxiety (b = -2.17; CI = -4.71; -0.37). We found no associations between COVID-19-specific variables and social and mental health in the psychiatric and pediatric groups.

### Atmosphere at home before and during COVID-19

Children from the psychiatric and pediatric samples reported no differences in atmosphere at home between before and during the COVID-19 lockdown, whereas children in the general population reported a worse atmosphere (ΔM = -3.13, p < 0.001) at home during the COVID-19 lockdown. All parents reported a negative change in atmosphere (psychiatric sample ΔM = -2.76, pediatric sample ΔM = -3.38, and general population sample ΔM = -4.62; all ps < 0.05; see Table 4). Furthermore, we investigated whether changes in atmosphere differed between the samples. The ANCOVA indicated no significant differences between the samples for children (p =.055, η^2^ = .005) and parents (p = .062, η^2^ = .005), corrected for age and gender.

**Table 4.** Atmosphere at home as experienced by children and parents.

|  | Atmosphere<br>before COVID-19 | Atmosphere<br>during COVID-19 | Change in<br>Atmosphere | *p* * |
| --- | --- | --- | --- | --- |
|  | *M* | *M* | *M* |  |
| Child/adolescent psychiatric sample (N=249) |  |  |  |  |
| Children | 72.9 | 72.4 | -0.6 | .675 |
| Parents | 74.1 | 71.4 | -2.8 | **.013** |
| Pediatric sample (N=90) |  |  |  |  |
| Children | 86.3 | 84.3 | -2.0 | .241 |
| Parents | 87.1 | 83.8 | -3.4 | **.028** |
| General population sample (N=844) |  |  |  |  |
| Children | 81.4 | 78.2 | -3.1 | **< .001** |
| Parents | 81.0 | 76.4 | -4.6 | **< .001** |

**Note.** \**P*-value of the paired sample T-test. A *p*-value <.05 indicates significant differences, shown in bold.
Parents answered the questions “How did you experience the atmosphere at home before the Corona regulations?” and “How do you experience the atmosphere at home now?” Children answered the questions: “How did you experience atmosphere at home before the schools were closed?” and “How do you experience the atmosphere at home now?” Answers were given on a visual analogue scale (range 0-100).

### Impact of the COVID-19-regulations on daily life

In general, children and adolescents from the vulnerable samples reported similar experiences with the COVID-19-regulations as the general population. Most children (86-92%) indicated that the COVID-19-regulations negatively impacted their daily life. The main problems they experienced (≥5% of the children per group) were missing contact with friends (∼42%), not being allowed to go to school (∼22%), missing freedom (∼16%), not being allowed to participate in sports (∼14%), missing joyful activities such as birthdays, shopping, day trips, and (graduation) parties (∼13%), and missing extended family (especially grandparents; ∼8%). However, there were some small differences between the experiences of the samples. Only children and adolescents from the pediatric and psychiatric sample reported difficulties with keeping social distance (e.g., hard to keep distance in daily life, not allowed to hug, and getting warnings from other people), whereas the general population sample was the only group that mentioned boredom due to the COVID-19-regulations. In addition, only children from the psychiatric sample indicated that the (continually changing) COVID-19-measures made them angry, sad, or insecure. A minority of children reported that their daily life was not affected by COVID-19-regulations (5-11%) or reported that the COVID-19-regulations positively influenced their daily life (e.g., ‘I actually like the measures, less social contact and clear rules’, ‘I like to be at home’; 3-5%). See Table 5 for an overview.

**Table 5.**
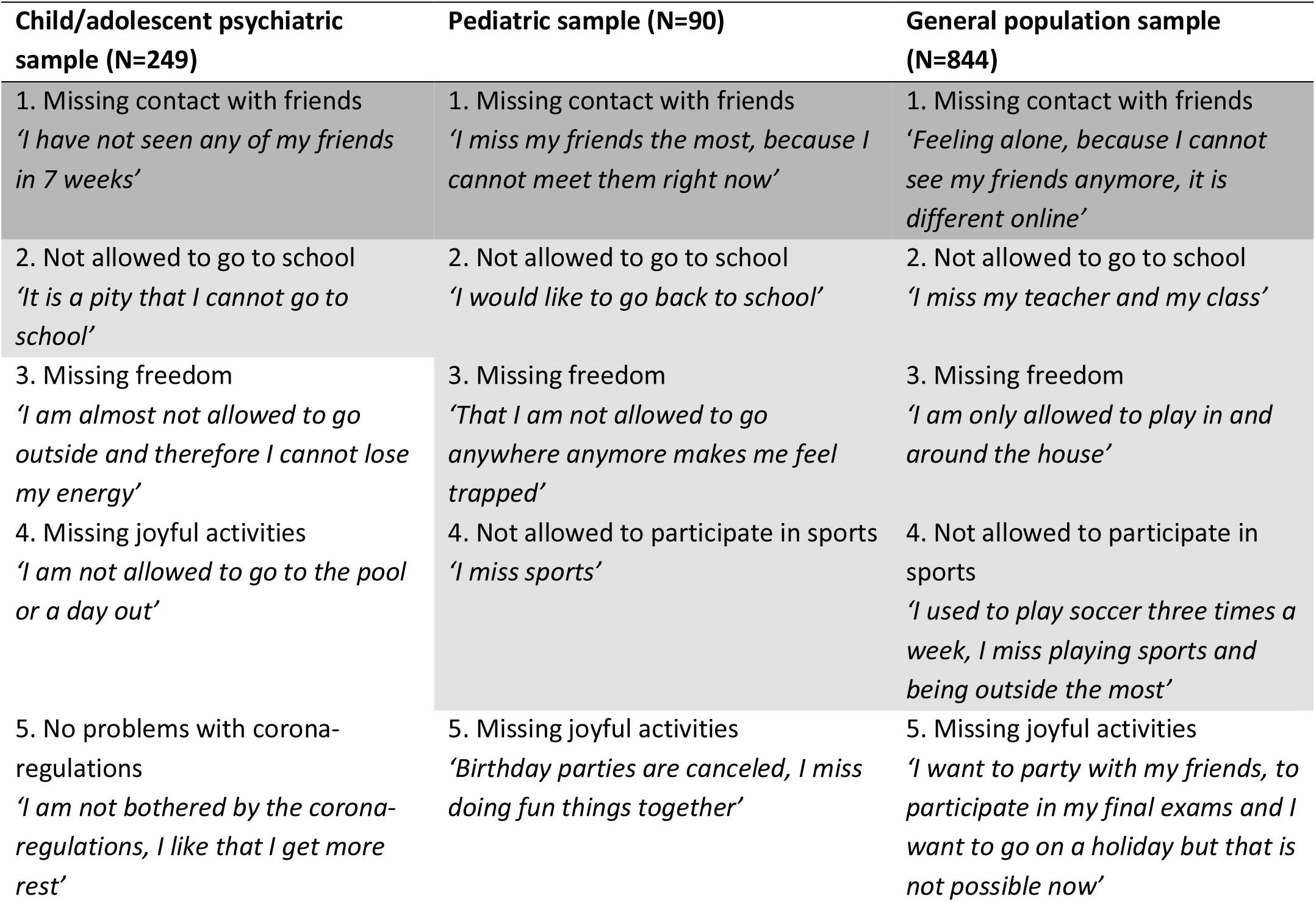
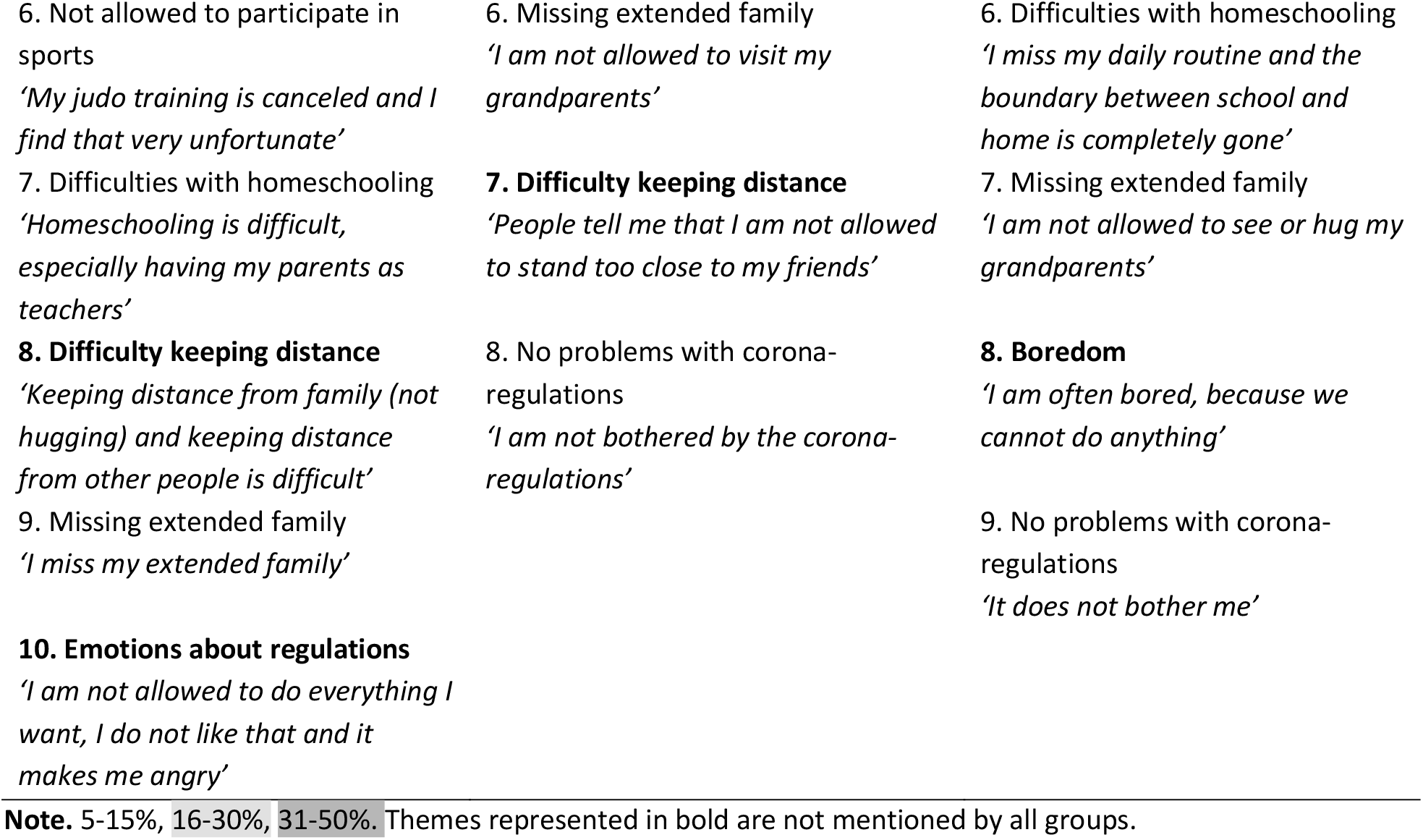
Overview of the most mentioned themes (≥5% of the children per sample) in response to the open question ‘How are the corona-regulations for you?’ per sample, ranked by the number of times mentioned (most to least)

## Discussion

In this study, we compared the mental and social health during the first Dutch COVID-19 pandemic lockdown in the spring of 2020 of children and adolescents with pre-existing psychiatric and somatic problems to a general population sample. We assessed global health, peer relationships, anxiety, depressive symptoms, anger, and sleep-related impairment. Mental and social health were worst for children with pre-existing psychiatric problems, whereas children from the pediatric sample showed the most favorable results.

As expected, children with pre-existing psychiatric problems showed most problems in mental and social health. The closing of schools and other services during the COVID-19 pandemic lockdown likely affects this group of children especially since these are places where vulnerable children often seek help first ^35^. Furthermore, access to professional mental health care substantially changed during the COVID-19 pandemic lockdown. Mental health care facilities were partly disrupted and had to rapidly adapt their services, while being perceived as having a higher threshold for help seeking, due to fear of infection. Additionally, people may have felt that mental problems are less important when the overall medical system is under high pressure because of COVID-19. Personal contact often changed to online therapy, of which the effects have not yet been tested thoroughly ^36^. Although online therapy can be an effective treatment, during the pandemic online therapy is sometimes provided out of necessity rather than choice. In addition, online therapy might increase the risk of losing contact with particular vulnerable families ^16^. Furthermore, existing feelings of anxiety might increase in response to the threat of close relatives being infected with COVID-19 or becoming infected oneself. The finding that children with pre-existing problems living in the north of the Netherlands showed the least anxiety problems might hint to this, since in that part of the country almost no reported COVID-19 cases were present at the time. Still, these interpretations are made with caution since based on the current data we cannot conclude that the COVID-19 lockdown plays a significant role in the higher levels of mental and social problems in the psychiatric population. The only data that clearly hint at this notion derive from the qualitative part of our study showing that only children from the psychiatric sample indicated that the (continually changing) COVID-19-measures made them angry, sad, or insecure.

In contrast to the psychiatric sample, children and adolescents from the pediatric sample showed the least problems on all mental and social health scales. As this population generally suffers from more psychosocial problems than the general population ^11,12^, this suggests they are least negatively affected by the COVID-19 pandemic lockdown regulations. A possible explanation for this finding may be that these children are more often confronted with stressful events that directly affect their physical health and restraints in daily life due to the management of their disease. As a result, children may have developed more adaptive coping strategies ^21,22^. In addition, one could speculate that the COVID-19 pandemic lockdown induced changes in daily life might be less invasive for some of these children as home schooling and carefully taking care of one’s physical health may be part of life already ^21^. Likewise, they may be more used to having online contact with friends, which may explain their higher reported Peer Relationships.

Concerning COVID-19-specific variables, a negative change in work situation of the parents and having an infected friend or relative seemed to contribute to mental and social health problems in the general population ^10^, but not in our samples of children and adolescents with pre-existing mental or somatic problems. It is possible that these children are less affected by such situations because they are more accustomed to dealing with stressful events and thereby have become more resilient to them. Since the qualitative part of this study suggested that the COVID-19-restrictions itself do affect mental health in the psychiatric population, it is important to examine additional COVID-19 related measures in the future.

Limitations of our study include low response rates and a potential selection bias of our samples, which limit extrapolation of our results. In particular, the pediatric sample was relatively small. Additionally, due to the cross-sectional design of this study, we cannot assess what factors have impacted on the *change* in mental and social health of children and adolescents due to the COVID-19 pandemic lockdown. Finally, there may be individual differences in how children respond to the lockdown. The literature so far hints at a potential diverse range of effects that the COVID-19 pandemic has on children and adolescents with pre-existing vulnerabilities, ranging from an increase in mental health problems to reduction of stress, loss of social pressure, and improved well-being ^20^. For future research, we recommend studying this variability more in depth.

This study contributes to ‘a call for action for mental health science’ by Holmes et al. ^37^ who argue that this type of research should be one of the main priorities in research during the COVID-19 pandemic. Our findings present descriptive data that compare mental and social health between vulnerable groups of children and adolescents and a general population. Future studies need to examine additional factors related to the COVID-19 pandemic lockdown and should apply longitudinal designs which will enable tracking the course of mental and social health when the COVID-19 lockdown regulations continue or return in the future.

## Supporting information

STROBE checklist

## Data Availability

Data are available upon reasonable request.

